# Electrochemical detection of myeloperoxidase (MPO) in blood plasma with surface-modified electroless nickel immersion gold (ENIG) printed circuit board (PCB) electrodes

**DOI:** 10.1101/2023.09.03.23295003

**Authors:** Ruchira Nandeshwar, Siddharth Tallur

**Affiliations:** Department of Electrical Engineering, Indian Institute of Technology Bombay, Mumbai 400076, MH India

**Keywords:** Myeloperoxidase, ENIG PCB, methylene blue, AuNP electrodeposition, cardiovascular disease

## Abstract

Printed circuit board (PCB) based biosensors have often utilized hard gold electroplating, that nullifies the cost advantages of this technology as compared to screen printed electrodes. Electroless nickel immersion gold (ENIG) is a popular gold deposition process widely used in PCB manufacturing, but vulnerable to pinhole defects and large surface roughness, which compromises biosensor performance. In this work, we present a method to address these challenges through electrodeposition of methylene blue (MB) to cover surface defects and improve electroactivity of ENIG PCB electrodes. We also demonstrate a process to realize in situ synthesis of gold nanoparticles (AuNPs) using acid-functionalized multi-walled carbon nanotubes (MWCNTs) as scaffold, that are used to immobilize antibody for the target molecule (myeloperoxidase: MPO, early warning biomarker for cardiovascular diseases) through a modified cysteamine/gluteraldehyde based process. The processing steps on the electrode surface are developed in a manner that do not compromise the integrity of the electrode, resulting in repeatable and reliable performance of the sensors. Further, we demonstrate a cost-effective microfluidic packaging process to integrate a capillary pump driven microfluidic channel on the PCB electrode for seamless introduction of samples for testing. We demonstrate the ability of the sensor to distinguish clinically abnormal concentrations of MPO from normal concentrations through extensive characterization using spiked serum and blood plasma samples, with a limit of detection of 0.202 ng*/*mL.

## I. INTRODUCTION

According to WHO, cardiovascular diseases (CVDs) are the leading cause of death due to non-communicable diseases, representing more than 30 % of global deaths [1]. Most CVDs can be prevented by addressing behavioral risk factors such as tobacco use, unhealthy diet and obesity, physical inactivity and harmful use of alcohol. It is important to detect CVDs as early as possible for effective disease management. There are various pathology-lab based blood tests to measure cholesterol levels and biomarkers of CVDs such as the gold standard cardiac troponin I, myoglobin, C-reactive protein (CRP), B-type natriuretic peptides (BNP), N-terminal prohormone BNP (NT-proBNP) etc. [2]. Monitoring these parameters, as well as measuring auxiliary parameters through ECG and echocardiography requires rapid access to a well-equipped lab/clinic and skilled technicians and doctors to perform and evaluate these tests. The associated economic and social costs are major obstacles to access for low-income and below-poverty line sections of populations in large, developing and lower-middle income countries (LMICs), which contribute more than three quarters of deaths due to CVDs [1]. The concentration of the biomarkers mentioned above elevates in the bloodstream at the onset of or immediately following myocardial infarction (commonly known as heart attack). Therefore, they are useful for diagnosing heart attacks, but cannot be leveraged as early warning biomarkers for detecting vulnerability to CVDs [2]. Additionally, researchers have found some early-stage markers which are released in the blood at the start of vascular inflammation. Myeloperoxidase (MPO) is one such marker which is released early in the blood stream before myocardial infarction [3], and has been established as an important biomarker to predict heart health [4, 5].

While at-home monitoring of blood glucose levels with low-cost point-of-care monitoring devices is playing an increasing role in management of diabetes, similar technology for monitoring of CVD biomarkers is missing. Some reports of electrochemical biosensors for MPO have been reported in literature. These biosensors use substrates such as glassy carbon electrodes [6, 7], glass [8] and screen printed electrodes [9–11]. However, such sensors are not directly capable of direct integration with electronic instrumentation, require custom fabrication processes for custom bonding techniques to integrate fluidic capabilities resulting in additional manufacturing costs, and do not meet the robustness requirement desired of point of care sensors. Recently, PCB electrodes and lab-on-PCB systems have gained attention being cost effective, easy to manufacture, compatible with electronic systems, and robust [12–17] (see detailed reviews by Shamkhalichenar et al. [18] and Moschou et al. [19]). To the best of our knowledge, PCB based biosensors for cardiovascular diseases have been reported in literature for only one CVD biomarker (cardiac troponin I) [20, 21].

Among various type of surface finish options available for PCBs [22], the hard gold electroplating process is widely used for lab-on-PCB biosensors despite being very expensive, due to protection against corrosion of copper tracks on the PCBs offered by the thicker gold film atop the copper and nickel layers. Hard gold finish also offers more uniform electroactive surface area and reduction of pinholes, thus improving sensitivity, repeatability and reliability of the sensors [17]. Reports of low-cost processes such as electroless nickel immersion gold (ENIG) finish PCB electrode based biosensors also employ additional gold electroplating [17, 23]. We recently demonstrated an ENIG finish PCB based immunosensor to detect SARS-CoV-2 spike protein in artificial saliva [24], wherein we reported methods to partially overcome several challenges associated with using ENIG finish PCB electrodes for electrochemical biosensors, e.g. corrosion of electrodes due to pinhole defects in the thin gold layer, and large surface roughness of the gold layer which poses difficulties in uniform deposition of antibodies. The most important challenges which remained unaddressed were the large electrode-to-electrode variation in performance observed due to surface non-uniformity, and limited sensitivity due to lack of nanoscale dimension features on the electrode surface. In this work, we present a method to address these challenges through electrodeposition of a redox active dye, methylene blue (MB), on the electrode surface along with in situ synthesis of gold nanoparticles (AuNPs) using acid-functionalized multi-walled carbon nanotubes (MWCNTs) as scaffold. The AuNPs are used to immobilize antibody for the target molecule (MPO) through a modified cysteamine/gluteraldehyde based process. All processing steps on the electrode surface are developed in a manner that do not compromise the integrity of the electrode, and the processes thus developed result in repeatable and reliable performance of the sensors, with no observations of the above-mentioned challenges. Further, we demonstrate a cost-effective microfluidic packaging process to integrate a capillary pump driven microfluidic channel on the PCB electrode for seamless introduction of samples for testing. We demonstrate the ability of the sensor to distinguish clinically abnormal concentrations of MPO from normal concentrations through extensive characterization using spiked serum and blood plasma samples, with a limit of detection of 0.202 ng*/*mL. An illustration of the workflow for utilization of the sensor is shown in Figure 1(a). Details of methods followed for surface modification of PCB electrodes, sample preparation, experimental protocols, results and discussion are presented in following sections.

**FIG. 1.**
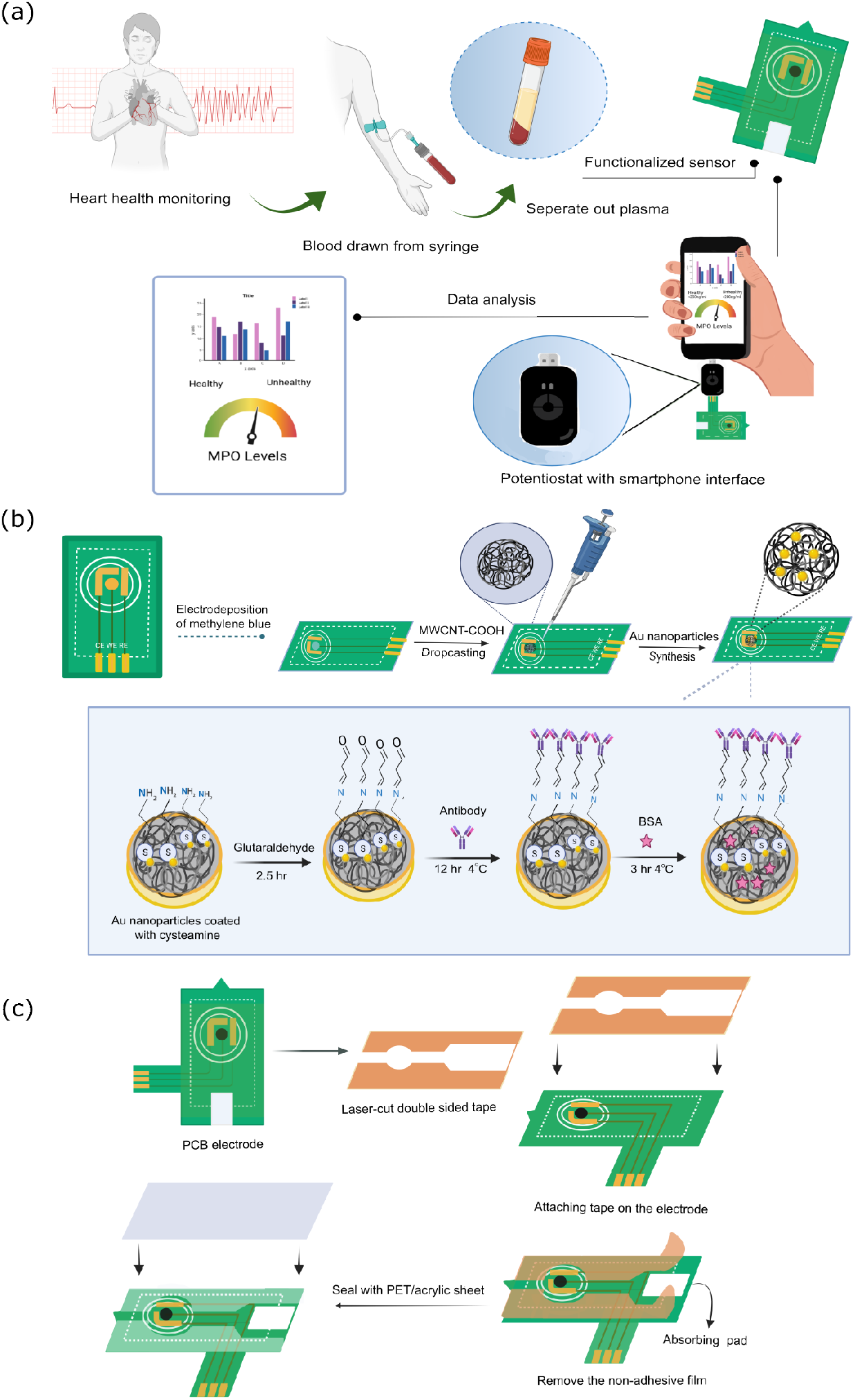
(a) Ilustration of sample collection and testing with functionalized printed circuit board (PCB) sensor. The sensor is designed such that it can be connected with PalmSens Sensit Smart potentiostat. Schematic representations of: (b) process flow for functionalization and modification of PCB electrodes, (c) process of integration of microfluidic channel on PCB electrode.

## II. MATERIALS AND METHODS

### A. Materials

95–98 % sulfuric acid (H_2_SO_4_), 65–70 % nitric acid (HNO_3_), sodium sulfate (Na_2_SO_4_) (catalog no. 239313), tetrachloroauric (III) acid (HAuCl_4_·3 H_2_O) (520918), 98 % ethanol, isopropyl alcohol (IPA), phosphate buffer saline (PBS) tablets (P4417), human serum (H4522), cysteamine (M9768), glutaraldehyde (G6257), bovine serum albumin (BSA) (A9085), methylene blue (C_16_H_18_ClN_3_S) (66720), multi-walled carbon nanotubes (755125) and dimethylformamide (DMF) were procured from Sigma Aldrich. Lyophilized human MPO was procured from Planta Natural Products (9003-99-0). MPO polyclonal antibodies (DOM00001G) and human MPO ELISA kit (BMS2038INST) were purchased from Thermo Fisher Scientific. Hydrophilized polytetrafluoroethylene (PTFE) filter membranes (47 mm diameter and 0.2 μm pore size) were procured from Axiva Sichem Pvt. Ltd. Ammonium hydroxide (30 % NH_4_OH) was purchased from Vetec and hydrogen peroxide (30 % H_2_O_2_) was procured from EMPARTA. Potassium ferrocyanide (K_4_[Fe(CN)_6_] *·* 3H_2_O) was used as redox probe. Venous blood sample was collected from a volunteer and stored in EDTA vacutainer at IIT Bombay hospital. All dilutions were prepared in DI water unless specified otherwise.

### B. PCB electrode design and cleaning process

PCB electrodes were designed using Autodesk EAGLE electronic design automation (EDA) tool. The design of the PCB electrode, with 3 mm diameter working electrode, is based on the design for SARS-CoV-2 immunosensor reported in our previous work [24]. These electrodes were manufactured with a conventional electroless nickel immersion gold (ENIG) plating process (Circuit Systems (India) Ltd.) These PCBs have a thin gold layer (thickness less than 100 nm) atop the nickel and copper layers on the PCB, and therefore significantly inexpensive (≈INR ₹50 per PCB electrode) as compared to PCBs with electroplated gold. These PCBs were cleaned with RCA-1 cleaning method. The PCBs were first wiped with IPA, and then dipped in ethanolacetone-DI water solution and sonicated in a bath sonicator for 15 min at 40 °C. The PCBs were then rinsed in DI water and wiped with a lint-free cloth, and dipped in 1 : 1 : 6 ammonium hydroxide-hydrogen peroxide-DI water solution and sonicated for 10 min at 40 °C. The PCBs were rinsed thoroughly in DI water and dried and stored at room temperature.

### C. Methylene blue coating on PCB

Various studies have been reported on polymerization of MB onto electrode surfaces [25–27]. In this work, MB is not polymerized but only coated on the working electrode surface. To coat MB on the PCB electrode surface, 80 μL of 2 mM MB solution was drop-casted on the PCB such that all three electrodes on the PCB (reference, workign and counter electrodes) were covered with MB. Cyclic voltammetry (CV) was performed for voltage range from −0.5 V to 0.5 V for 10 cycles at 50 mV*/*s scan rate. The excess MB was then rinsed with DI water, and the PCBs were air-dried at room temperature.

### D. Acid functionalization of multi-walled carbon nanotubes and coating on PCB electrodes

Multi-walled carbon nanotubes procured from Sigma Aldrich (*>*8 % carboxylic acid functionalized, 9.5 nm average diameter × 1.5 μm length) were subjected to additional acid functionalization to further increase the percentage of carboxylic functional groups. The as-procured and further functionalized multi-walled carbon nanotubes are referred to as MWCNT and MWCNT-F subsequently in the manuscript. The acid functionalization process was based on methods conventionally reported in literature e.g., Jun et al. [28], Wepasnick et al. [29] and Blanchard et al. [30]. 50 mg of MWC-NTs were added to 27 mL of 18.4 M H_2_SO_4_ and 9 mL of 15.8 M HNO_3_. The suspension was sonicated in bath sonicator for 2.5 h at 40 °C. After sonication, the suspension was diluted in DI water. The samples were then filtered with the help of a vacuum filter assembly using PTFE filter membranes. The functionalized MWCNT-F samples were then rinsed with DI water till neutral pH was achieved, and then dried in an oven at 60 °C for 12 h and stored in a glass vial at 4 °C. The MWCNT-F samples were dispersed in dimethylformamide (DMF) to obtain a concentration of 1 mg*/*mL and sonicated in a bath sonicator for 2 h to achieve uniform dispersion. To coat the PCB electrode, 4 μL of the nanotubes dispersed in DMF were drop-casted only on the working electrode (WE) and dried for 12 h at room temperature. PCB electrodes were similarly prepared with MWCNT, for evaluating the efficacy of acid functionalization by comparing with the MWCNT-F coated electrodes. Visibly uniform coating of MWCNT and MWCNT-F was obtained on the WE.

### E. Electrodeposition of gold nanoparticles (AuNPs) on MWCNT-F coated PCBs

AuNP deposition process reported by Rabai et al. [31] was modified for ENIG PCBs. Solution of 0.1 mM HAuCl_4_ and 0.2 M Na_2_SO_4_ was drop-casted on the MB/MWCNT-F PCB electrode such that it covered the reference, working and counter electrodes on the PCB. Chronoamperometry was performed at a potential step of −0.8 V for 200 s, 400 s and 500 s to electrodeposit AuNPs on the acid functionalized multi-walled carbon nanotubes on the working electrode. The PCBs were then rinsed with DI water and dried at room temperature.

### F. Immobilization of MPO antibody on PCB

The MB/MWCNT-F/AuNP coated working electrode on the PCB was modified by drop-casting 20 μL of 10 mM cysteamine (mixed in ethanol) at room temperature. Excess cysteamine was rinsed with DI water. 12 μL of 2.5 % of glutaraldehyde mixed in DI water was then drop-casted on the cystamine-coated working electrode, and the PCBs were left undisturbed at 4 °C for 2.5 h. Excess glutaraldehyde was rinsed with DI water. 10 μL of 10 μg*/*mL MPO antibody was then drop-casted on the working electrode, following which the PCB electrodes were left undisturbed at 4 °C for 12 h. The PCB electrodes were thereafter rinsed with PBS buffer and 10 μL of 1 % bovine serum albumin (BSA) was drop-casted on the antibody-coated working electrode at 4 °C for 3 h, to prevent non-specific binding of MPO and other proteins present in the sample on the electrode. Excess BSA was rinsed with PBS buffer and the modified electrodes were stored at 4 °C. A schematic illustration of the electrode modification process is shown in Figure 1(b).

### G. Microfluidic packaging of PCB electrodes

Figure 1(c) shows an illustration of the process for integration of microfluidic channel on the PCB electrode for seamless and controlled introduction of test samples on the active area of the electrode. The microfluidic packaging of the sensor was realized using polyethylene terephthalate (PET) sheet and 3M double-sided tape. The double-sided tape was cut in the desired pattern with the help of a laser cutting machine (SIL Accucut 6090) and then aligned and stuck firmly on the PCB electrode. The channels were sealed with a PET sheet after adding an absorbent pad (cut-out piece of lint-free wipe). Final height of the channel was 350 μm. Detailed drawings are provided in supplementary information (Figure S1).

### H. Extraction of plasma from blood sample

Blood sample was collected from a volunteer and stored in BD K2-EDTA vacutainer to prevent clotting. The sample was centrifuged at 2000 g for 15 min to separate plasma from whole blood. The plasma was collected in 1.5 ml centrifuge tubes in separate aliquots and was stored at −20 °C until required for further use. One of the aliquots was used on the same day, for establishing baseline concentration of MPO using ELISA. The ELISA kit (Thermo Fisher Scientific, BMS2038INST) was operated as per guidelines provided by the manufacturer.

### I. Sample preparation and characterization

#### 1. Physical testing

Material coatings on PCB electrodes were characterized using Raman spectroscope, atomic force microscope (AFM), X-ray photoelectron spectroscope (XPS), Fourier transform infrared (FTIR) spectroscopy and scanning electron microscope (SEM). Raman spectra were recorded using LabRAM HR Evolution-RAMAN and AFM images were recorded on Asylum/Oxford Instruments, model no. MFP3D Origin. Raman spectra of MB drop-casted on Si wafer were used as a control for comparing Raman spectra of MB coated on PCB. XPS spectra were recorded with Kratos Analytical, model AXIS Supra. FTIR spectra of MWCNT and MWCNT-F were recorded with 3000 Hyperion Microscope with Vertex 80 FTIR System (Bruker, Germany). Pellets of MWCNT and MWCNT-F were made with KBr. For obtaining SEM micrographs, MWCNT and MWCNT-F were drop-casted on the working electrode and dried at room temperature. AuNPs were electrodeposited on the PCB as described in section II E. Imaging was performed using a field emission scanning electron microscope JEOL JSM-7600F (JEOL Ltd., Japan). Dimensions of features in the micrographs were analyzed using ImageJ software.

#### 2. Electrochemical characterization of sensor response

For electrochemical characterization (cyclic voltammetry - CV, and differential pulse voltammetry - DPV) of the sensor, 10 mM potassium ferrocyanide in 0.1 M KCl solution was used as redox probe. All measurements were performed using PalmSens Sensit Smart potentiostat and analyzed using PalmSens PSTrace software. The impact of MB coating on the PCB was studied by comparing CV voltammograms obtained before and after coating of MB on PCB, for voltage range −0.2 V to 0.5 V and 100 mV*/*s scan rate. The impact of MWCNT-F deposition, AuNP growth, and immobilization of antibody on the electrode was studied by first obtaining the sensor dose-response curve with MPO spiked in PBS buffer. DPV voltammograms were used to study the dose-response of MPO, for voltage range −0.2 V to 0.3 V and 50 mV*/*s scan rate. Serial dilutions of MPO in PBS buffer were prepared to obtain final concentrations of 1 ng*/*mL, 5 ng*/*mL, 10 ng*/*mL, 20 ng*/*mL, 50 ng*/*mL, 100 ng*/*mL, 200 ng*/*mL and 300 ng*/*mL. For studying sensor response in the presence of a complex matrix such as human serum, MPO was spiked in commercially procured serum to obtain the following concentrations: 100 ng*/*mL, 200 ng*/*mL, 300 ng*/*mL and 500 ng*/*mL. These samples of MPO in serum were then diluted by a factor of 10 with PBS buffer to minimize bio-fouling of the sensor surface. A similar process was repeated with plasma separated from whole blood. The following concentrations of MPO were spiked in plasma, over and above the baseline MPO concentration present in the sample: 50 ng*/*mL, 100 ng*/*mL, 200 ng*/*mL and 300 ng*/*mL. These spiked plasma samples were then diluted by a factor of 10 in PBS buffer. For studying the impact of EDTA in the vacutainer on the sensor response, commercially procured serum without EDTA was compared with serum spiked with K2-EDTA, and these two sets of serum were spiked with several concentrations of MPO (50 ng*/*mL, 150 ng*/*mL and 300 ng*/*mL). Each sample was then diluted by a factor of 10 with PBS buffer before testing. Serum spiked with K2-EDTA had the same concentration of EDTA (1.8 mg*/*mL) as that present in a 6 mL BD vacutainer.

## III. RESULTS AND DISCUSSION

### A. Methylene blue coating reduces variability across PCB electrodes

Unlike electroplated gold, ENIG finish gold pads on low-cost PCBs have greater surface non-uniformity and defects such as black pad defects and pinhole defects formed during manufacturing. These defects can lead to corrosion of the PCB [32]. The problem is exacerbated by reduction of electroactivity due to ionic (e.g., P, Cu) and organic (e.g., fluxes, resins) impurities present on the PCB surface, that are also introduced during the manufacturing process [33]. In this work, we have focused on addressing issues such as surface non-uniformity, pinhole defects, corrosion and contamination due to organic impurities on the PCB electrodes. Studying the impact of ionic impurities is beyond the scope of this work, and could be investigated in future work. Organic impurities are removed from the electrode surface using RCA-1 cleaning method, as described in section II B. We have previously shown the efficacy of this method for improving the electroactivity of PCB electrodes [24]. Electrode-to-electrode variability is one of the most important challenges in electrochemical biosensors, that affects screen printed electrodes (SPEs), and is particularly worse for PCB-based biosensors. The use of a thick electroplated gold layer in PCB-based electrochemical biosensors is primarily to ensure a low roughness surface with no defects, thus resulting in uniformity of antibody immobilization. In this work, we have explored electrodeposition to remediate the electrode-to-electrode variation in ENIG finish PCB electrodes. Electrodeposition and electropolymerization methods have been reported to offer higher uniformity than manual/machine-based approaches [34, 35]. Due to susceptibility of ENIG finish PCBs to corrosion on account of pinhole defects in the thin gold layer, such deposition steps should be performed without applying large potential difference across the electrodes, and without the use of corrosive chemicals such as strong acids or bases on the electrode. To accomplish this, we electrodeposited methylene blue dye diluted in DI water on the PCB electrode, without applying large voltages or using corrosive chemicals. Methylene blue dye is a phenothiazine dye and is used as a redox probe in many applications due to its electrochemical activity [25, 27, 36–39]. Typically, a high potential (*>*0.8 V) and low pH (5) are needed for the formation of cation radicals that initiate the polymerization process, and ensure stability of the polymer film [40]. Applying such high potentials and maintaining low pH is not possible with ENIG PCB electrodes due to the risk of corrosion. Therefore in this work, we electrodeposited MB on the electrodes at neutral pH (7) instead of performing electropolymerization. The detailed process of deposition is described in section II C.

Two electrodes each were coated with methylene blue through 5 CV cycles and 10 CV cycles with 2 mM MB. After performing the electrodeposition, voltammograms were recorded 15 times on each electrode by adding the redox probe (potassium ferrocyanide and KCl solution), to study the variability of the electrochemical response. These measurements were also performed on a control electrode, with no MB coating. The oxidation and reduction peak currents were measured for each CV voltammogram, and the results are shown as a bar graph of the average relative standard deviation (RSD) of CV repeated 15 times on these electrodes in Figure 2(a). Electrodes coated with 10 CV cycles of MB show 30 − fold reduction in RSD (approximately 1 %) as compared to control electrodes (approximately 30 %). By means of representation, Figure 2(b) shows voltammograms recorded during coating of MB through 10 successive CV cycles on one of these electrodes. No peak is observed for cation formation in these CV voltammograms, which confirms that there is no polymerization of MB. The oxidation peak current gradually increases with each CV cycle, indicating adsorption of more and more MB resulting in formation of a thin layer on the electrode surface. CV voltammograms obtained with potassium ferrocyanide and KCl on 3 electrodes coated with MB using 10 CV cycles are shown in Figure 2(c). The variability across the three electrodes reduced drastically due to electrodeposition of MB. The anodic and cathodic peak currents also increased signficantly, and the separation between the anodic and cathodic peak potentials also reduced from 100 mV to 45.5 mV after electrodeposition of MB, indicating an improvement in electroactivity and electrochemical stability of the electrode surface [41]. Electrodes coated with 10 CV cycles of MB were used for all further experiments reported in this manuscript.

**FIG. 2.**
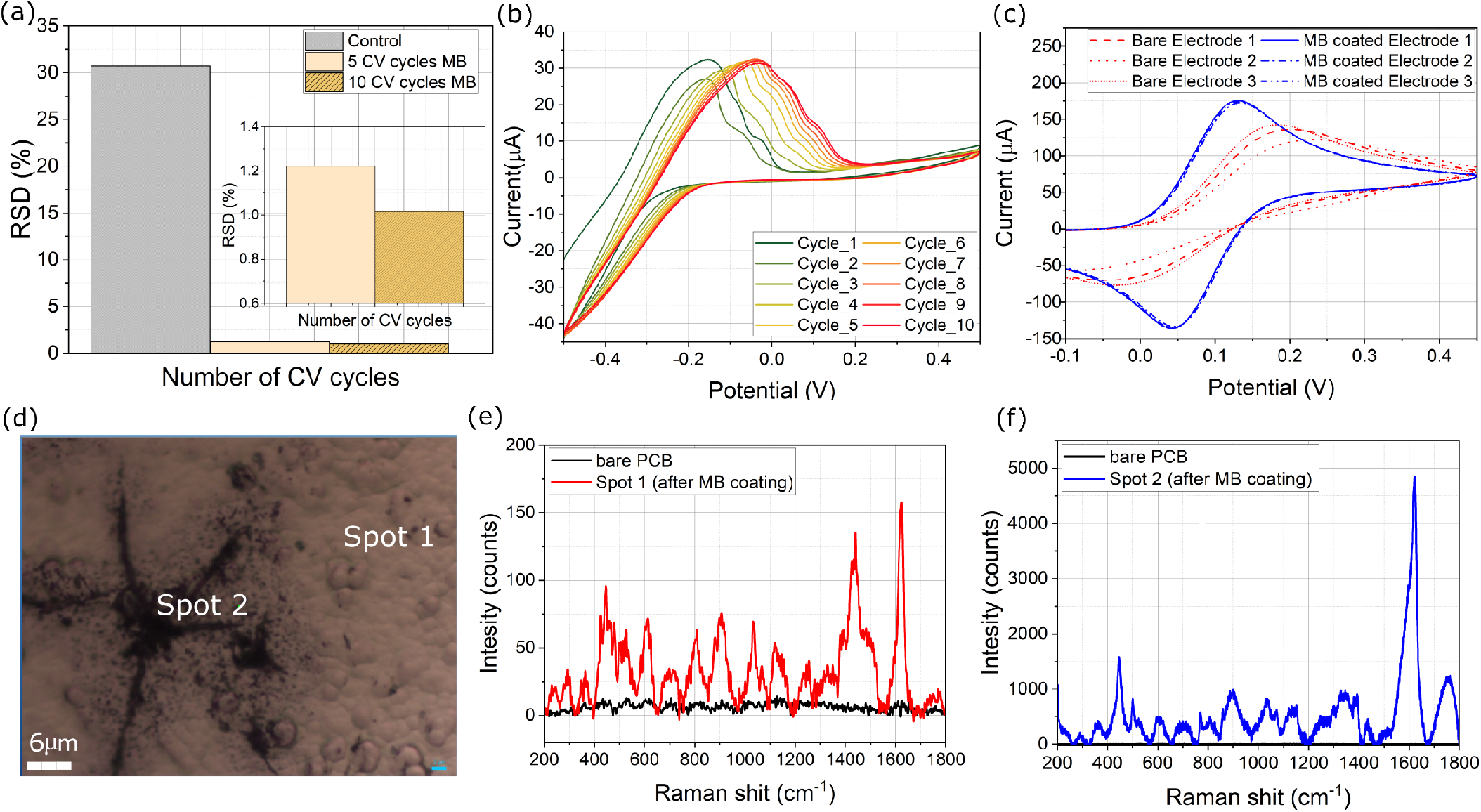
(a) Comparison of relative standard deviation (RSD) of oxidation and reduction peak of redox probe (repeated 15 times) on MB-coated electrodes following 5 and 10 CV cycles and, and a control electrode with no MB coating. (b) Voltammogram recorded during 10 CV cycles (voltage range −0.5 V to 0.5 V at 50 mV*/*s scan rate) for MB deposition on a PCB electrode. (c) CV voltammograms recorded on 3 PCB electrodes, before (bare electrode) and after electrodeposition of MB. (d) Optical micrograph of PCB surface with defects and pinholes, showing 2 spots where Raman spectra were recorded. (e) Raman spectra of bare PCB electrode (no MB coating) and spot 1 (after MB coating). At spot 1, the Raman intensity is very low which indicates that a thin layer of MB is adsorbed on the surface. (f) Raman spectra of bare PCB (no MB coating) and spot 2 (after MB coating). The spectrum recorded at spot 2 indicates deposition of multiple layers of MB.

To verify the presence of MB on the electrode surface, Raman spectroscopy was performed on the electrodes. The signature of MB in Raman spectra was determined by comparing Raman spectra obtained on a silicon wafer on spot coated with MB by drop-casting with an uncoated portion of the wafer (Figure S2 in supplementary information). This experiment was also used to investigate any differences in drop-casted and electrodeposited MB. Figure 2(d) shows a micrograph of two spots on PCB electrode coated with MB that were investigated through Raman spectroscopy. Spot 1 is on an area with no surface defect in the ENIG finish gold, while spot 2 lies on an area with a defect on the PCB. The Raman spectra measurements for these spots are shown in Figures 2(e) and (f) respectively, wherein they are compared to measurement obtained on a bare PCB i.e., with no MB coating. The characteristic peaks of MB seen on the silicon wafer coated with MB are also observed on the PCB electrodes: 445 cm^−1^, 498 cm^−1^, 480 cm^−1^, 605 cm^−1^, 762 cm^−1^, 1034 cm^−1^, 1072 cm^−1^, 1148 cm^−1^, 1301 cm^−1^, 1340 cm^−1^, 1390 cm^−1^, 1439 cm^−1^ and 1625 cm^−1^. This observation confirms that MB monomers get adsorbed on the PCB surface [42]. Peaks at 445 cm^−1^ and 498 cm^−1^ are attributed to skeletal deformation of C−N−C bond. The peak at 1340 cm^−1^ is due to symmetrical stretching of C−N bond and the peak at 1439 cm^−1^ is due to asymmetrical stretching of C−N bond. The peak at 1625 cm^−1^ is attributed for C−C ring stretching [43, 44]. While the Raman spectra confirm the adsorption of MB on both spots on the PCB electrode, it is interesting to observe that the intensity of the peaks recorded at spot 2 are significantly higher than those recorded at spot 1. This observation suggests that more MB monomers are deposited at the defect site, and therefore the surface becomes more uniform, thus explaining the reduction in electrode-to-electrode variability. This hypothesis is further corroborated by the observation that the peak at 480 cm^−1^ (corresponding to monolayer formation) is present only in spot 1 Raman spectra, while peaks at 445 cm^−1^ and 498 cm^−1^ (corresponding to multilayer formation) are observed at spot 2 [45]. The presence of these peaks also supports the claim that more MB is adsorbed at defect sites making the surface smooth. The impact of electrodeposition of MB on the surface roughness of the PCB electrode was characterized with AFM. The average RMS surface roughness of PCB electrodes reduced from 97.54 nm to 70.80 nm after coating. AFM micrographs are included in supplementary information (Figure S3).

The Raman spectra peaks at 356 cm^−1^ and 1439 cm^−1^ might provide additional insights on the nature of MB-Au binding. The peak at 1439 cm^−1^ is attributed to asymmetrical stretching of C−N bond, and could indicate that nitrogen in MB is interacting with the gold surface on the PCB. This hypothesis is also supported by the presence of a small peak at 356 cm^−1^, which is attributed to N−Au binding [46]. This peak is only present in the spectra obtained on PCB electrodes and is not present in Raman spectra obtained on the silicon wafer. The evidence of N−Au binding is also supported by XPS spectra shown in Figure 3. The peaks at 83.3 eV and 87.01 eV peaks in Figure 3(b) correspond to Au 4f_7*/*2_ and Au 4f_5*/*2_ of the ENIG finish gold PCB electrode [47]. Figure 3(c) shows a peak for N 1s, with deconvoluted peaks at 396.4 eV, 399.3 eV, 401.28 eV that correspond to C−N/ −C−NH, −C=N− and −NH respectively [27, 48]. The peak for O 1s shown in Figure 3(d) with deconvoluted peaks at 530.89 eV for Me−OX(metal-oxide), 531.7 eV for O=C, and 533.35 eV for O−C [49]. Figure 3(e) shows peak for C 1s and deconvoluted peaks at 284.43 eV for C−C/C−H, 285.5 eV for C−OH/C−N and 287.5 eV for C=O [27, 49, 50]. These peaks confirm the absorption of MB on the PCB electrode and the sulfur peak was not observed in the XPS spectra. This indicates that MB might bind to the PCB surface through Au−N binding.

**FIG. 3.**
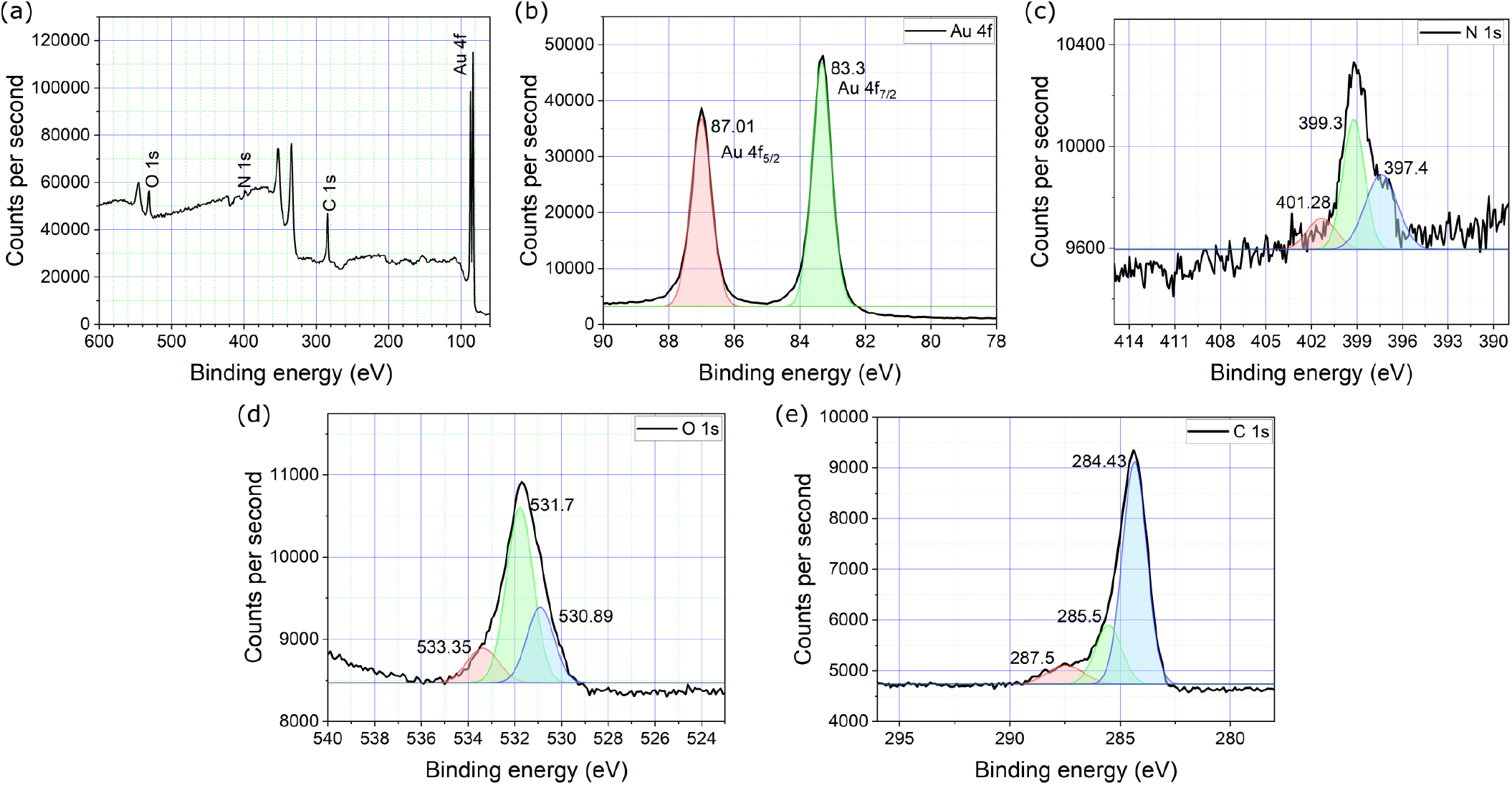
X-ray photoelectron spectroscopy (XPS) spectra: (a) Survey spectrum to identify prominent peaks. (b)-(e): Core-level and deconvoluted XPS peaks for (b) Au 4f, (c) N 1s, (d) O 1s, and (e) C 1s.

### B. AuNP deposition enhances the electrochemical activity of PCB electrodes

Incorporation of nanoparticles on the sensor surface improves sensitivity and better electron transfer rate [51, 52]. Nanomaterials can be synthesized separately and then introduced on the sensor surface, or the nanomaterial can be grown in situ on the electrode surface by methods such as electrodeposition, electrospinning, sputtering, chemical vapor deposition, lithography etc. [53]. All reported methods have their advantages and limitations, and the method that is most compatible with the sensor substrate should be followed for any sensor development. In the case of ENIG PCB electrodes, corrosion is a major challenge, and therefore processes requiring high temperature or pressure, large potentials, and strong acids or bases are not compatible with ENIG PCB electrodes. In this work, we focused on the use of AuNPs on ENIG finish PCB electrodes. depositing nanomaterials and reducing electrode-to-electrode variability therefore we used electrodeposition of gold nanoparticles (AuNP) on the ENIG finish PCB. AuNPs are extensively used in biosensing applications owing to their excellent biocompatibility and ability to bind to various biomolecules [54]. Electrodeposition of AuNPs has been reported on a variety of substrates in literature, however to the best of our knowledge, this work is the first report of electrodeposition of AuNP on ENIG finish PCBs.

We used acid-functionalized MWCNTs as scaffolds to grow AuNP on MB-coated PCB electrodes. Acid-functionalized MWCNTs enhance the hydrophilicity of the substrate and form a porous membrane, thereby reducing bio-fouling of the electrode surface [55, 56]. Details of AuNP electrodeposition process are described in section II E. The AuNPs were grown in a neutral supporting electrolyte (Na_2_SO_4_) to prevent corrosion of PCB electrodes [57]. Acid functionalization of MWCNTs also reduced agglomeration when the nanotubes were dispersed in DMF. Well-dispersed oxidized-MWCNTs have more sites to initiate the nucleation process of metal nanoparticle deposition. The process for acid functionalization of MWCNT with H_2_SO_4_ and HNO_3_ is explained in section II D. FTIR spectra shown in Figure 4(a) confirm the carboxylic acid functionalization of MWCNT. The peaks for C−O stretch at 1162 cm^−1^, C−O in the range 1637 cm^−1^ to 1712 cm^−1^, C−H stretch at 2925 cm^−1^ and 2854 cm^−1^, O−H stretch at 3448 cm^−1^ show higher intensity for MWCNT-F as compared to MWCNT [28]. The peak at 1567 cm^−1^ is a signature of carboxylate anion [58]. Note that the as-procured MWCNTs had *>*8 % carboxylic acid functionalization. SEM micrographs of MWCNT and MWCNT-F (after functionalization) are shown in Figure 4(b). The images were analyzed using ImageJ software to determine the diameters of the nanotubes. A slight increase in diameter is noticed for MWCNT-F as compared to MWCNT without acid functionalization, as shown in Figure 4(c). The increase may be explained by the introduction of carboxylic groups and defects at the side walls of the nanotubes [59, 60]. Additional supporting figures based on ImageJ analysis are provided in supplementary information (Figure S4).

**FIG. 4.**
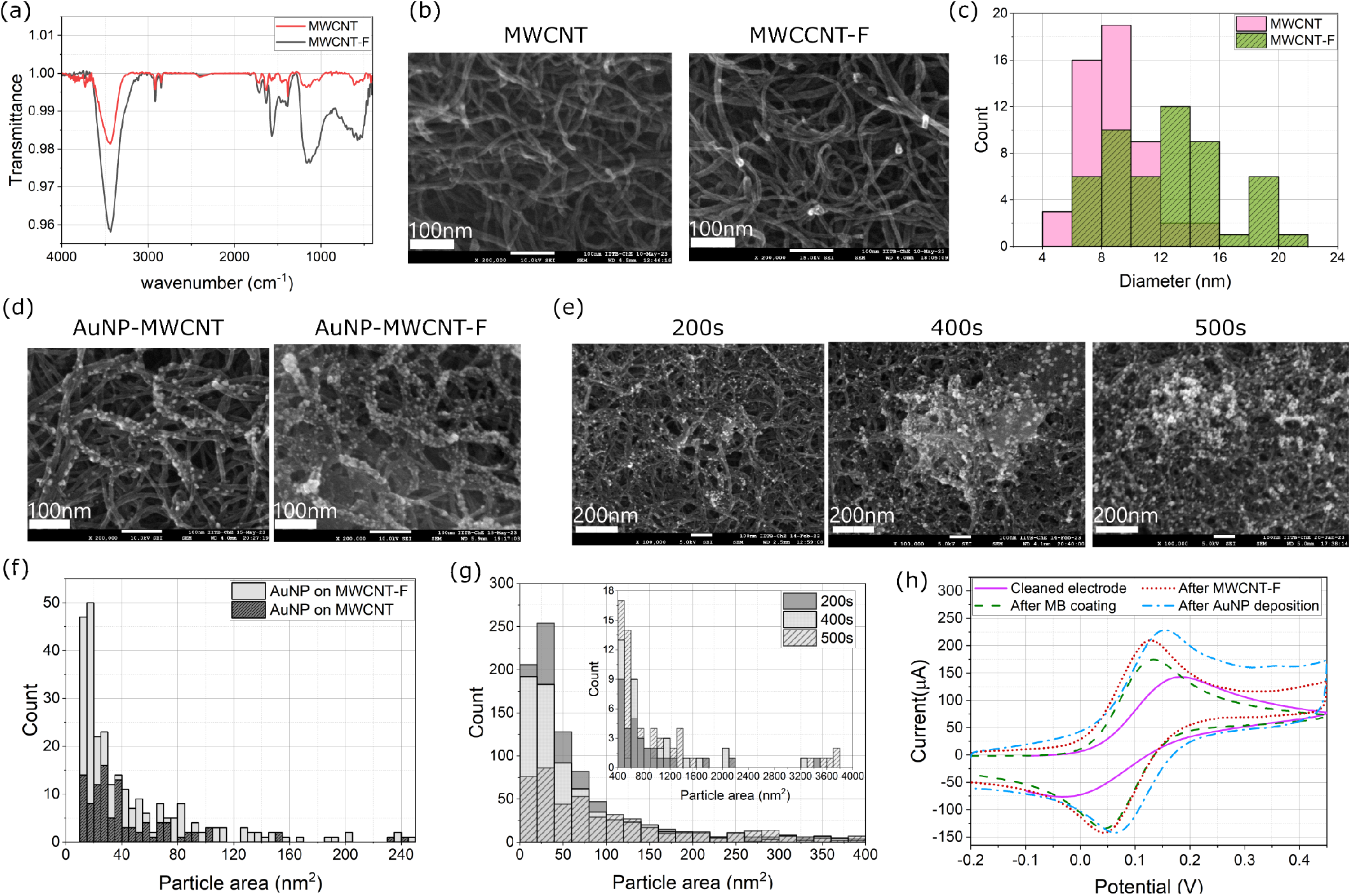
(a) Fourier transform infrared (FTIR) spectra of as-procured MWCNTs (label: MWCNT) and acid functionalized MWCNTs (MWCNT-F). (b) SEM micrographs of MWCNT and MWCNT-F drop-casted on PCB electrodes. (c) Distribution of diameters of MWCNTs before and after acid functionalization. (d) SEM micrographs of AuNPs electrodeposited on MWCNT and MWCNT-F. (e) SEM micrographs of AuNPs electrodeposited on MWCNT-F using chronoamperometry for 200 s, 400 s and 500 s. (f) Distribution of particle area (obtained through ImageJ analysis of SEM micrographs) of AuNPs electrodeposited on MWCNT and MWCNT-F. (g) Distribution of particle area (obtained through ImageJ analysis of SEM micrographs) of AuNPs electrodeposited on MWCNT-F using chronoamperometry for 200 s, 400 s and 500 s. (h) CV voltamogramms recorded on a PCB electrode after each modification step in the following order: (1) cleaned electrode, (2) after MB coating, (3) after (dropcasting) MWCNT-F, (4) after AuNP deposition.

AuNPs were electrodeposited on MWCNT and MWCNT-F as mentioned in section II E through chronoamperometry. for 200 s. SEM micrographs of AuNPs grown with chronoamperometry for 200 s on MWCNT and MWCNT-F are shown in Figure 4(d). Figure 4(e) shows SEM micrographs of AuNPs grown on MWCNT-F for varying durations of chronoamperometry (200 s, 400 s and 500 s). The size and density of the AuNPs in Figure 4(d) was analyzed, and the results are shown in Figure 4(f). From the histograms, it can be observed that while the particle sizes (represented as area) are comparable for MWCNT and MWCNT-F, the density of particles (i.e., number of counts) is significantly higher for MWCNT-F. This is expected, since MWCNT-F has more functional groups than MWCNT, which leads to more AuNP deposition. Au^3+^ ions get reduced to Au at these defect sites and initiate nucleation. Distribution of AuNP particle size (area in the SEM micrograph) for various durations of chronoamperometry shown in Figure 4(g) indicate that the spread of the distribution increases with increasing duration of chronoamperometry. This is due to formation of clusters of AuNP, as seen in the micrographs shown in Figure 4(e). These bigger clusters increase the surface-to-volume ratio of features on the sensor surface and can aid in immobilization of higher density of antibodies. Therefore, AuNPs were deposited on MWCNT-F for 500 s to obtain maximum sensitivity of the sensor. The time duration was not increased beyond 500 s to avoid corrosion of the PCB electrodes. Additional images based on ImageJ analysis to estimate particle sizes are included in supplementary information (Figure S4). After each modification step, CV voltammogram was recorded using the redox probe, and the results are shown in Figure 4(h). The peak current increases after each modification step, confirming successful deposition of all materials on the sensor surface.

### C. Clinically relevant concentrations of MPO can be detected with modified PCB electrodes

Details of coating PCB/MB/MWCNT-F/AuNP electrode with cysteamine, glutaraldehyde, MPO antibody and BSA are described in section II F. BSA was added to prevent non-specific binding of other analytes on the electrode. Cysteamine binds to AuNPs through thiol linkages and the amino groups of cysteamine are protonated and positively charged at pH = 7, which attracts more negatively charged redox probe ions to the electrode surface. This should lead to an increase in CV current [61], and was confirmed by recording CV voltammograms before and after coating cysteamine on the electrode (Figure S5 in supplementary information). Detection of MPO using these functionalized PCB electrodes was performed by recording DPV voltammograms and observing changes in the peak current corresponding to the redox potential for potassium ferrocyanide. MPO dilutions were prepared in PBS as per the process described in section II I 2. The samples (10 μL) were introduced serially on the electrode in increasing order of concentration. After allowing the MPO to incubate on the electrode for 15 min and bind with the antibody, the electrode was washed with DI water and then 80 μL of the redox probe was added on the electrode for performing DPV measurement. Thereafter, the electrodes were again rinsed with DI water, and the process was repeated with the sample containing next higher concentration of MPO. DPV voltammograms for MPO concentrations in PBS recorded on one electrode are shown in Figure 5(a). ‘Blank’ denotes DPV recorded before incubating any samples containing MPO on the PCB electrode. The DPV peak current decreases with increasing MPO concentration as more antigen binds with antibodies on the electrode surface blocking the diffusion of electrons. To obtain the limit of detection (LOD) and to check the variability across electrodes, the testing process was repeated on three different electrodes. The DPV peak currents readings for all MPO concentrations were normalized with respect to DPV peak current reading for 1 ng*/*mL MPO concentration, and the average and standard deviation (*σ*) were calculated. Results shown in Figure 5(b) indicate very low variability across electrodes, with a log-linear regression fit *R*^2^ = 0.99, and estimated LOD (3.3*σ/S*) = 0.2 ng*/*mL, where *S* denotes sensitivity of the PCB electrode sensors. Additional supporting results are included in supplementary information (Figure S6).

**FIG. 5.**
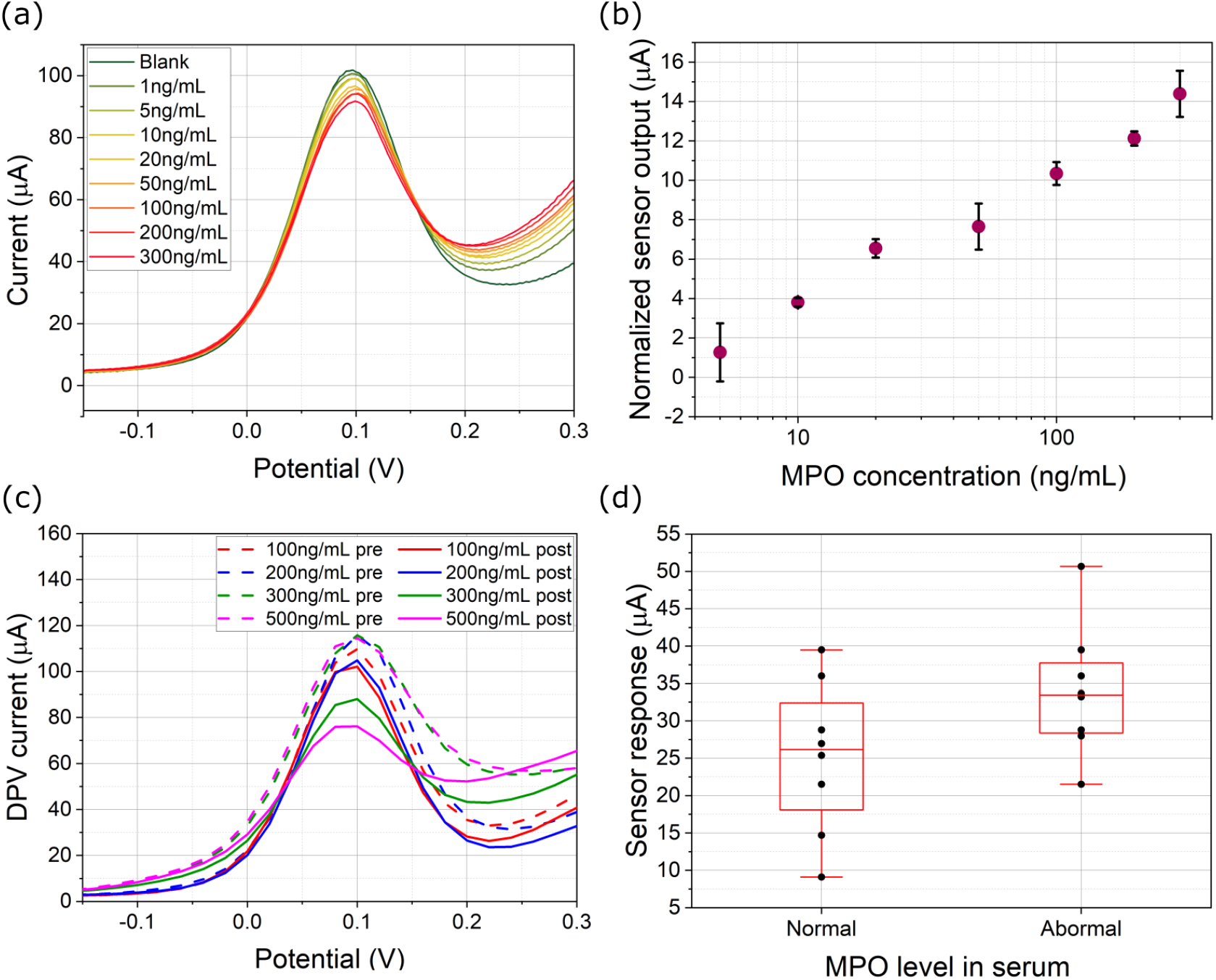
(a) DPV voltammograms recorded for various MPO concentrations diluted in PBS buffer tested on a modified PCB electrode. The samples were tested in increasing order of concentration. ‘Blank’ denotes reading obtained before dispensing any MPO samples on the electrode. (b) Dose-response curve of the sensor for MPO in PBS. The data is normalized with respect to sensor response for 1 ng*/*mL MPO in PBS. (c) DPV voltammograms recorded for MPO spiked in 10X diluted human serum samples. ‘pre’: background reading before dispensing MPO sample, ‘post’: reading obtained after adding MPO sample (incubation time: 15 min). (d) Sensor response for abnormal and normal MPO concentrations spiked in commercially procured human serum samples. The sensor response is the change in peak current level from ‘pre’ to ‘post’.

For a point-of-care/point-of-use biosensor, it is very important that the device can detect clinically relevant antigen concentrations in real samples. Before testing the sensor with human venous blood sample, the modified sensor was tested with commercially procured human serum. The commercially procured human serum is a suitably complex matrix to test the sensor response and study the efficacy of the surface modification process for the anti-fouling of the electrode. Serum samples spiked with different MPO concentrations were prepared as described in section II I 2. Before incubating MPO spiked serum sample on the electrode, the background DPV reading of each modified electrode was recorded by drop casting the redox probe on the modified electrode.

Following this step, the electrode was rinsed with DI water, and 10 μL of the MPO spiked serum sample was drop-casted on the modified working electrode on the PCB, and left undisturbed for 15 min to allow antigen-antibody binding to occur. The excess sample was rinsed with DI water and 80 μL of the redox probe was drop casted on the electrode to measure DPV current. The sensor response for a particular MPO concentration was recorded as the change in DPV peak after adding the sample from the DPV peak current in the background reading obtained on the same electrode. Each MPO concentration was tested on three different electrodes. The electrodes were not reused i.e., only two readings were taken on each electrode: background reading, and reading with serum sample spiked with a particular concentration of MPO. Representative DPV voltammograms for one electrode each for each concentration of MPO and corresponding background readings are shown in Figure 5(c). The graph shows that the change in the peak current of the sample (post) with respect to the background peak current (pre) increases with an increase in MPO concentration. The change is comparatively much lower for 100 ng*/*mL and 200 ng*/*mL MPO concentration in serum as compared to 300 ng*/*mL and 500 ng*/*mL. The MPO concentrations were classified into two categories i.e, abnormal MPO levels (above 200 ng*/*mL i.e., 300 ng*/*mL and 500 ng*/*mL) and normal MPO level (100 ng*/*mL and 200 ng*/*mL) as per reported MPO levels in literature [62]. The sensor is able to distinguish between abnormal and normal levels of MPO in commercially procured human serum as illustrated in Figure 5(d) with p-value 0.08, calculated using JMP® software. Additional supporting results are included in supplementary information (Figure S5).

Another important aspect of point-of-care/point-of-use devices is the ease of sample testing and automation of sample processing. To achieve this goal, we incorporated microfluidic packaging into the PCB electrodes, the details of which are described in section II G. The microfluidic packaging is capillary-driven and no additional pump is needed to push the sample inside the channel. The design has two regions: (i) testing region (circular area) that exposes all three electrodes (RE: reference electrode, WE: working electrode, CE: counter electrode) to the sample, and (ii) capillary pump region at the other end containing absorbing pad, to collect excess liquid. For realizing a scalable process compatible with batch manufacturing, we used inexpensive materials such as PET sheets and double-sided tape, machined with laser cutting instead of using PDMS mold. The PCB electrodes were first functionalized, and the background peak current was recorded on the modified electrodes before packaging them. Figure 6(a) shows the packaged PCB sensor connected to PalmSens Sensit Smart potentiostat, which is connected to a mobile phone for data acquisition and visualization. Figure 6(b) shows photographs of a modified PCB electrode before (top) and after incorporation of the microfluidic packaging (bottom).

**FIG. 6.**
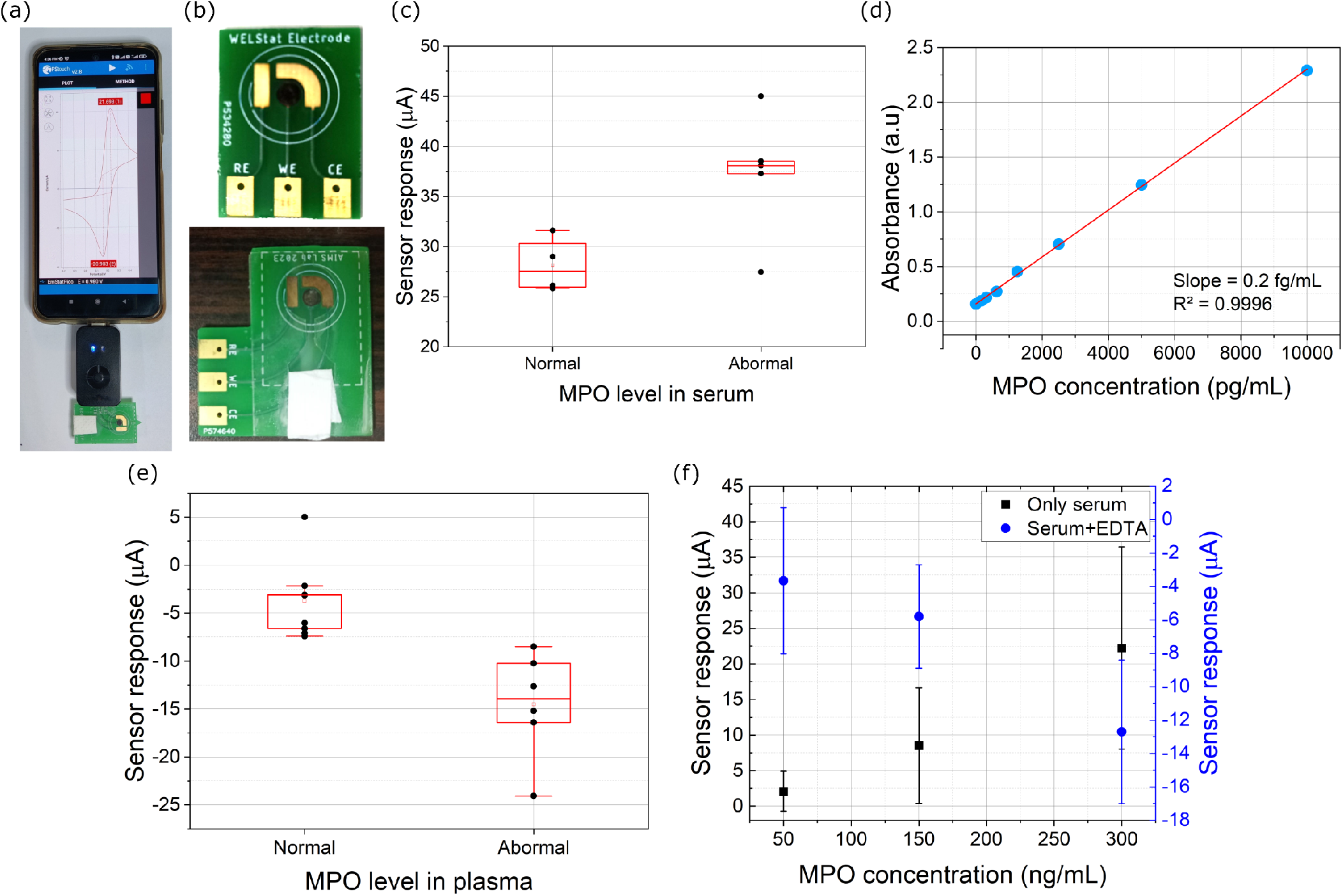
(a) PCB electrode with microfluidic packaging connected to PalmSens Sensit Smart potentiostat, and data visualized using smartphone application. (b) Top: modified working electrode without microfluidic packaging. Bottom: modified electrode with microfluidic packaging. An absorbing pad is added to the capillary pump section on the electrode. (c) Sensor response for normal (100 ng*/*mL and 200 ng*/*mL) and abnormal (300 ng*/*mL and 500 ng*/*mL) MPO levels spiked in serum samples, tested on electrodes with microfluidic packaging. (d) Standard curve of MPO ELISA assay. (e) Sensor response for normal (73.21 ng*/*mL, 123.21 ng*/*mL and 173.21 ng*/*mL) and abnormal (273.21 ng*/*mL and 373.21 ng*/*mL) MPO levels spiked in plasma samples. (f) Comparison of sensor response for serum samples spiked with MPO, with and without added EDTA, showing reversal in trend due to presence of EDTA.

The packaged sensors were tested with MPO spiked in commercially procured human serum samples. Samples were prepared as per the process described in section II I 2. 40 μL of serum sample spiked with MPO was introduced via the microfluidic channel, which is sufficient to cover the active area of the electrodes. After 15 min of incubation, the test sample was pushed out by adding enough redox probe instead of rinsing the test sample with DI water. This step was performed in such a manner that all the sample in the channel was absorbed by the absorbing pad, thus ensuring that excess sample was removed from the testing region. Then 30 μL of redox probe is again added to the sensor strip and DPV is recorded. Each packaged sensor was used only once i.e., only two readings were taken on each sensor: background reading before packaging, and reading with serum sample spiked with a particular concentration of MPO after packaging. The difference between these two values is noted as the sensor response, and the results are shown in Figure 6(c). The packaged sensors can detect abnormal levels of MPO in human serum (above 200 ng*/*mL) with a p-value of 0.028. Additional supporting results are included in supplementary information (Figure S5). Additional photographs and videos of testing of the microfluidic packages are included in supplementary information (Figure S6, and supplementary videos SV1 and SV2).

The modified sensor was also tested with a blood sample. The sample collection and plasma extraction process is described in section II H. The baseline level of MPO in the sample was determined by ELISA, performed on the same day as the sample was processed. The standard curve obtained from the ELISA kit is shown in Figure 6(d), and the baseline MPO concentration in the plasma separated from the blood sample was calculated to be 73.21 ng*/*mL. The following concentrations of MPO were spiked in plasma, over and above the baseline MPO concentration present in the sample: 50 ng*/*mL, 100 ng*/*mL, 200 ng*/*mL and 300 ng*/*mL. Therefore, the MPO concentrations in the samples thus prepared were: 73.21 ng*/*mL (no additional MPO was spiked), 123.21 ng*/*mL, 173.21 ng*/*mL, 273.21 ng*/*mL and 373.21 ng*/*mL. These samples were tested on modified electrodes with the same testing protocol as followed during the testing of MPO spiked in commercially procured serum samples. Each MPO concentration was tested on 3 different electrodes and the electrodes were not reused. The change in DPV peak current as compared to background reading was recorded for each electrode, and the results are shown in Figure 6(e). The sensors were able to distinguish between abnormal (273.21 ng*/*mL and 373.21 ng*/*mL) and normal levels (73.21 ng*/*mL, 123.21 ng*/*mL and 173.21 ng*/*mL) of MPO in the plasma samples with p-value of 0.003. However, unlike readings reported for commercially procured serum samples spiked with MPO and MPO dilutions prepared in PBS, the sensor response for MPO in plasma samples is negative i.e., the DPV peak current for the background was lesser than that obtained after adding plasma sample containing MPO on the sensor. Additional supporting results are included in the supplementary information (Figure S5).

This reversal of trend in the current response could be explained by the fact that the blood sample was collected and stored in a vacutainer containing EDTA, which could affect the sensor response. Scheffer et al. has suggested the use of EDTA-plasma samples for such experiments, as heparin-plasma and blood clotting agents lead to in vitro release of MPO from activated leukocytes [63], also supported by observations reported by Gerasimov et al. [64]. MPO is heme-containing peroxidase, with an Fe^3+^ ion in its structure [65]. It has been established that EDTA has a strong affinity to Fe^3+^ ion, resulting in the formation of a redox-active Fe−EDTA complex [66, 67]. Therefore, it is possible that this Fe−EDTA complex is in close vicinity of the electrode surface due to MPO binding with the antibody immobilized on the electrode, resulting in interference of the electrochemical response. To test this hypothesis, commercially procured serum without EDTA was compared with serum spiked with 1.8 mg*/*mL K2-EDTA, and these two sets of serum were spiked with several concentrations of MPO (50 ng*/*mL, 150 ng*/*mL and 300 ng*/*mL). These samples were tested with modified PCB electrodes and the change in peak current for each sample was calculated. Figure 6(f) shows the change in peak current for various concentrations of MPO spiked in serum, with and without EDTA, which confirms the reversal of trend in the peak current observed with spiked plasma samples.

## IV. CONCLUSION AND FUTURE WORK

In this work, we used ENIG finish PCB electrodes to electrochemically detect myeloperoxidase an early warning biomarker for cardiovascular diseases. Typically, electroplated PCB electrodes with hard-gold plating are generally used in biosensing applications, since they are not vulnerable to several challenges that plague ENIG finish PCB electrodes, such as susceptibility to corrosion, large variations in performance across multiple PCBs, presence of pinhole defects on the electrode etc. In this work, we have demonstrated an effective method to resolve these issues, through the electrodeposition of methylene blue on the electrodes.

The MB-coated electrodes demonstrated good repeatability, improved electroactivity, and reduced electrode-to-electrode variability. Raman spectroscopy revealed that the MB layer is not uniform and multiple MB layers get deposited near defect sites on the PCB. This is a significant insight toward understanding the mechanism of improving PCB biosensor performance using such coatings. XPS spectra revealed further insight into the mechanism of binding of MB with the gold surface on the electrode through Au−N binding. Although the characterization results support that MB binds to the PCB surface and covers the defects, more experiments, and detailed studies are needed to understand the exact nature of the interaction between MB and ENIG finish gold on the PCB electrode, which is beyond the scope of this manuscript and may be addressed in future work.

The sensor performance was enhanced through the electrodeposition of AuNPs on acid-functionalized MWCNTs on the electrode. The aforementioned challenges pertaining to ENIG finish PCBs also limit the applicability of conventional methods for in situ synthesis of nanoparticles on the electrodes. To the best of our knowledge, this work is the first report on in situ electrodeposition of AuNPs on ENIG PCB electrodes. To prevent corrosion of electrodes during deposition, we did not use acid as a supporting electrolyte and instead adapted the process to work with a neutral salt (Na_2_SO_4_). This process protected the electrode from corrosion and facilitated controlled and uniform growth of AuNPs, which would not be possible with drop-casting of presynthesized nanoparticles onto electrodes coated with MWCNTs. We also demonstrated the efficacy of acid functionalization of MWCNTs, which increased the percentage of carboxylic acid groups on the nanotubes thereby resulting in a higher density of AuNPs. The AuNPs increased the surface-to-volume ratio for immobilizing MPO antibodies, resulting in higher sensitivity. The LOD of the modified PCB electrode sensor calculated from the MPO dose-response curve was 0.202 ng*/*mL, which is suitable for a point-of-care biosensor, considering that the at-risk level of MPO concentration in blood is 200 ng*/*mL. A comparison of this work with other electrochemical sensors for MPO reported in the literature is provided in Table 1 in supplementary information. To automate the sample testing process, we incorporated a simple capillary-driven microfluidic channel onto the PCB electrode. The microfluidic packaging for the sensor was realized with inexpensive materials (3M double-sided tape and PET sheet) processed using a laser-cutting and engraving machine. We also successfully demonstrated the ability of the sensor to detect abnormal and normal levels of MPO in spiked serum and plasma samples. The origin of the reversal of trend in sensor response with increasing MPO concentration in spiked plasma samples was also successfully investigated and attributed to the presence of EDTA in the vacutainer used to collect and store the blood sample. The sensor performance could be further enhanced by using other sensitive techniques like electrochemical impedance spectroscopy (EIS), and developing an array of electrodes functionalized for different biomarkers of interest to realize a multiparameter sensor. This would also require addressing the cross-sensitivity of various functionalized electrodes to other biomarkers in the sample and the development of batch-compatible manufacturing processes. Further improvement in the anti-fouling performance could be investigated so that the sensor can be tested directly with undiluted blood samples, akin to at-home use glucometers.

**TABLE I.**
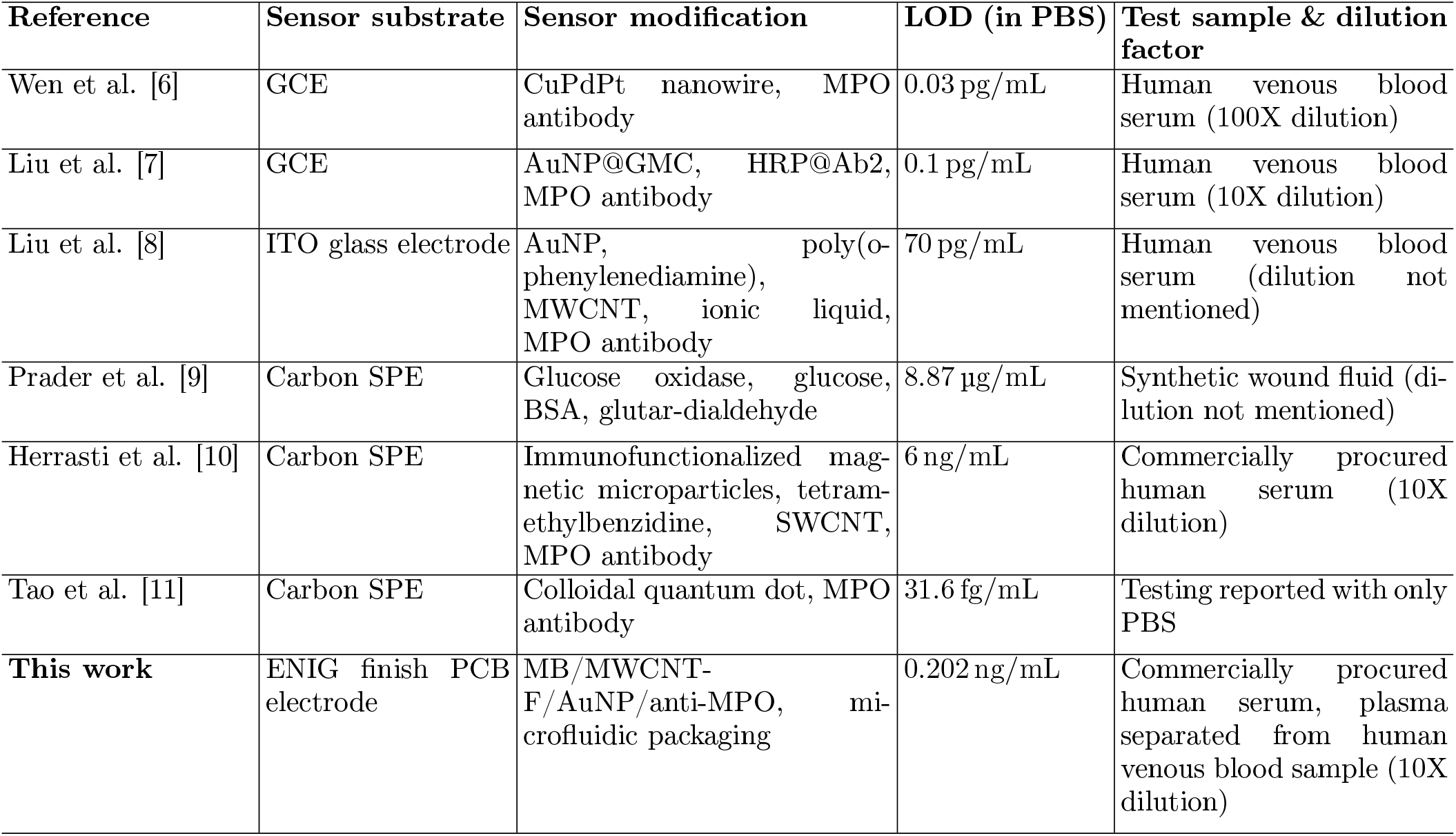
Summary of various electrochemical sensors reported in literature for detection of MPO. Glossary − LOD: limit of detection, GCE: glassy carbon electrode, SPE: screen printed electrode, ITO: indium tin oxide, SWCNT: single-walled carbon nanotube, GMC: graphitized mesoporous carbons, HRP: horseradish peroxidase

## Data Availability

All data produced in the present study are available upon reasonable request to the authors

## ACKNOWLEDGMENTS

R.N. acknowledges Ministry of Education (formerly Ministry of Human Resource Development), Government of India, for supporting her Ph.D. scholarship. This work was supported by a grant from the Gandhian Young Technological Innovation Award (GYTI) 2021 awarded to R.N. (project ID: BT/BIRAC/SITARE-GYTI0597) by Biotechnology Industry Research Assistance Council (BIRAC), a not-for-profit Public Sector Enterprise set up by Department of Biotechnology (DBT), Government of India. The authors thank Mr. M. Santosh Kumar, Ph.D. student at the Department of Biosciences and Bioengineering at IIT Bombay, for assistance in performing ELISA measurements; Prof. Debjani Paul at the Department of Biosciences and Bioengineering at IIT Bombay, for providing access to laboratory facilities for handling and processing blood samples; Ms. Riddhi Shah, B.Tech. student at Department of Mechanical Engineering at IIT Bombay, for assistance with preliminary design of microfluidic channels; and Ms. Soma Ghosh for help with preparing illustrations for Figure 1. The authors acknowledge various facilities at IIT Bombay for access to testing and sample characterization facilities: Sophisticated Analytical Instrument Facility (SAIF), IIT Bombay Nanofabrication Facility (IITBNF), Wadhwani Electronics Lab (WEL), and central facilities established through Industrial Research and Consultancy Centre (IRCC).

## ETHICS APPROVAL

Venous blood sample was collected from a volunteer and stored in EDTA vacutainer at IIT Bombay hospital, for some of the experiments reported in this study. All precautions were followed as approved by the Institute Biosafety Committee (IBSC) and Institute Ethics Committee (IEC, approved proposal ID: IITB-IEC/2021/021) at IIT Bombay.

## DATA AVAILABILITY

The data that support the findings of this study are available upon reasonable request from the authors.

## Supplementary Information

**FIG. S1.**
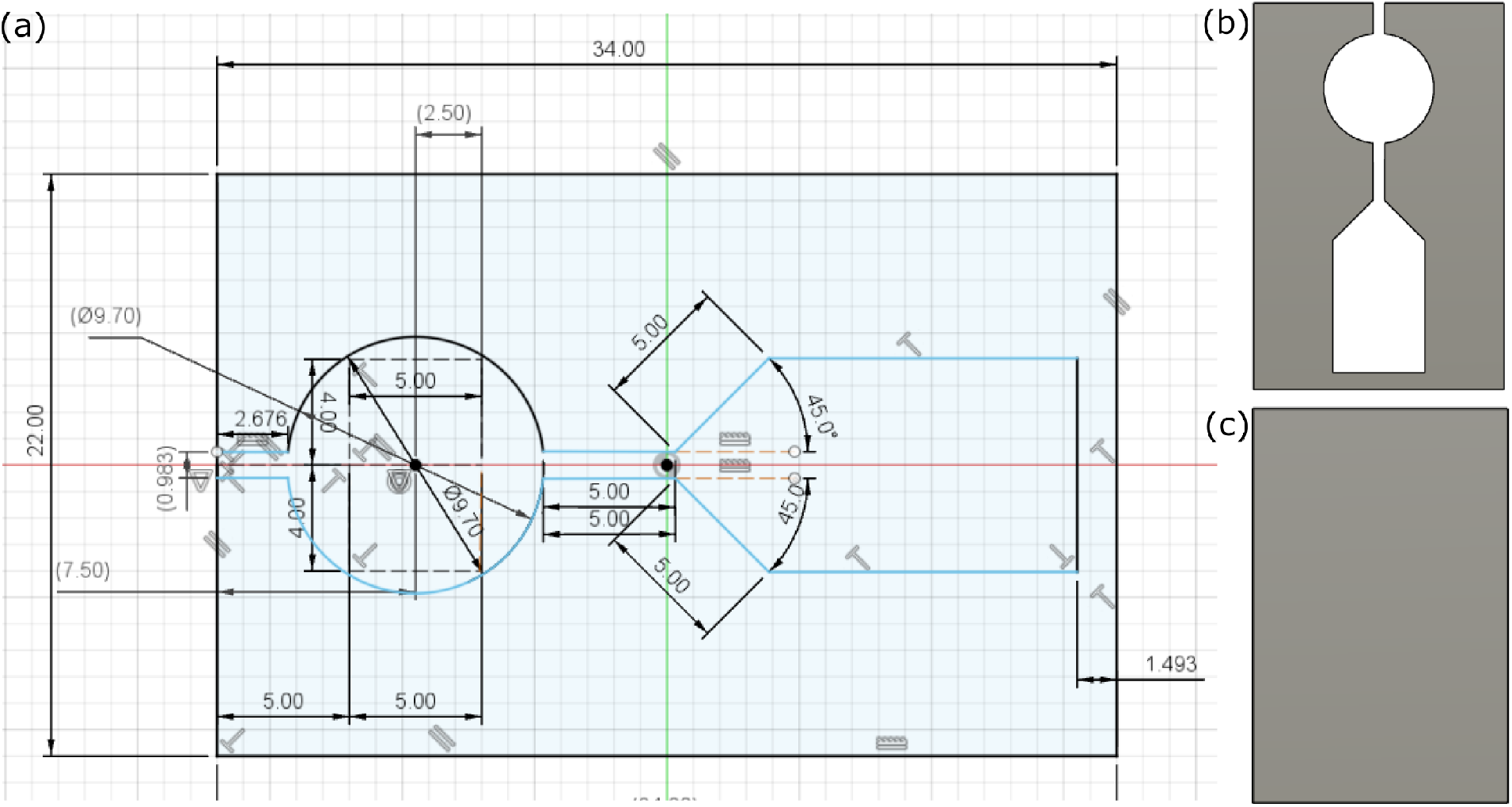
(a) Sketch of microfluidic packaging designed using Autodesk® Fusion 360™ software, highlighting relevant dimensions (mm). (b) 2D drawing of the double-sided 3M tape sketeched in panel (a). (c) 2D drawing of PET sheet used for encapsulation of the channel.

**FIG. S2.**
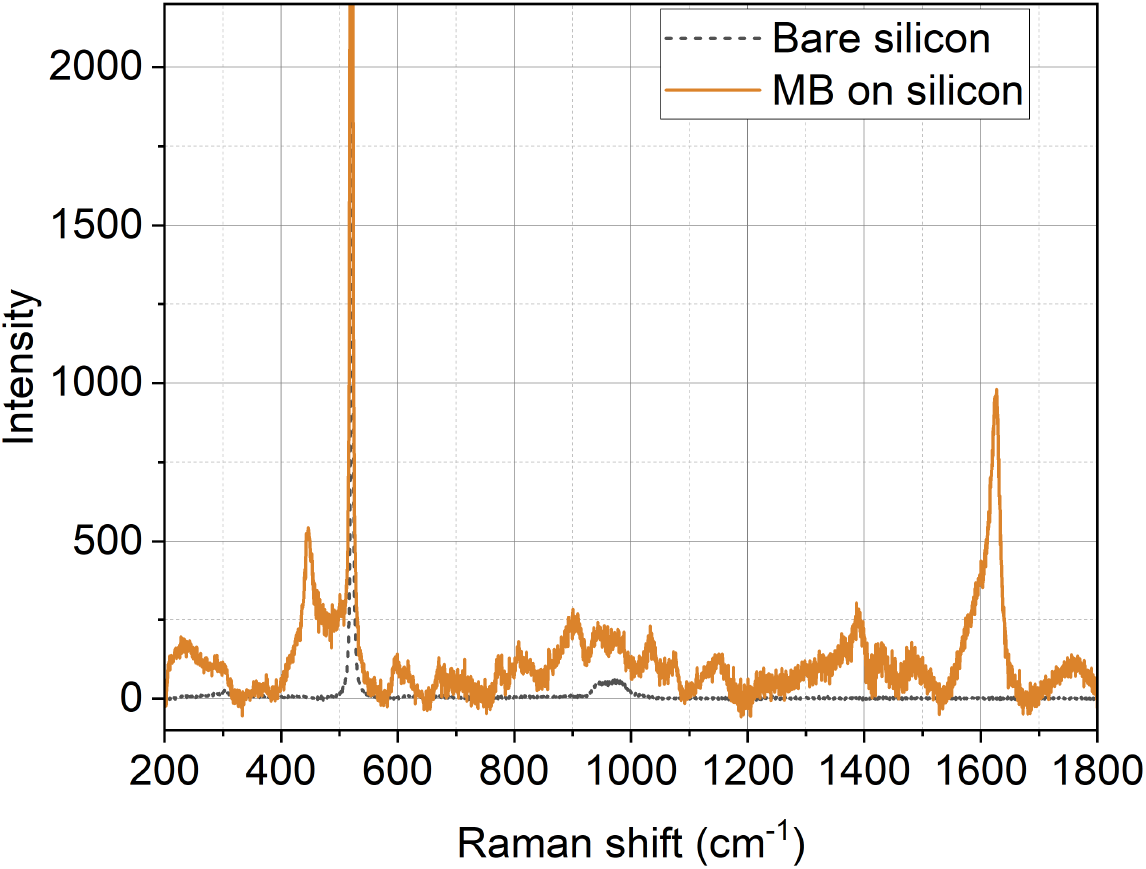
Raman spectra of methylene blue drop-casted on Si wafer.

**FIG. S3.**
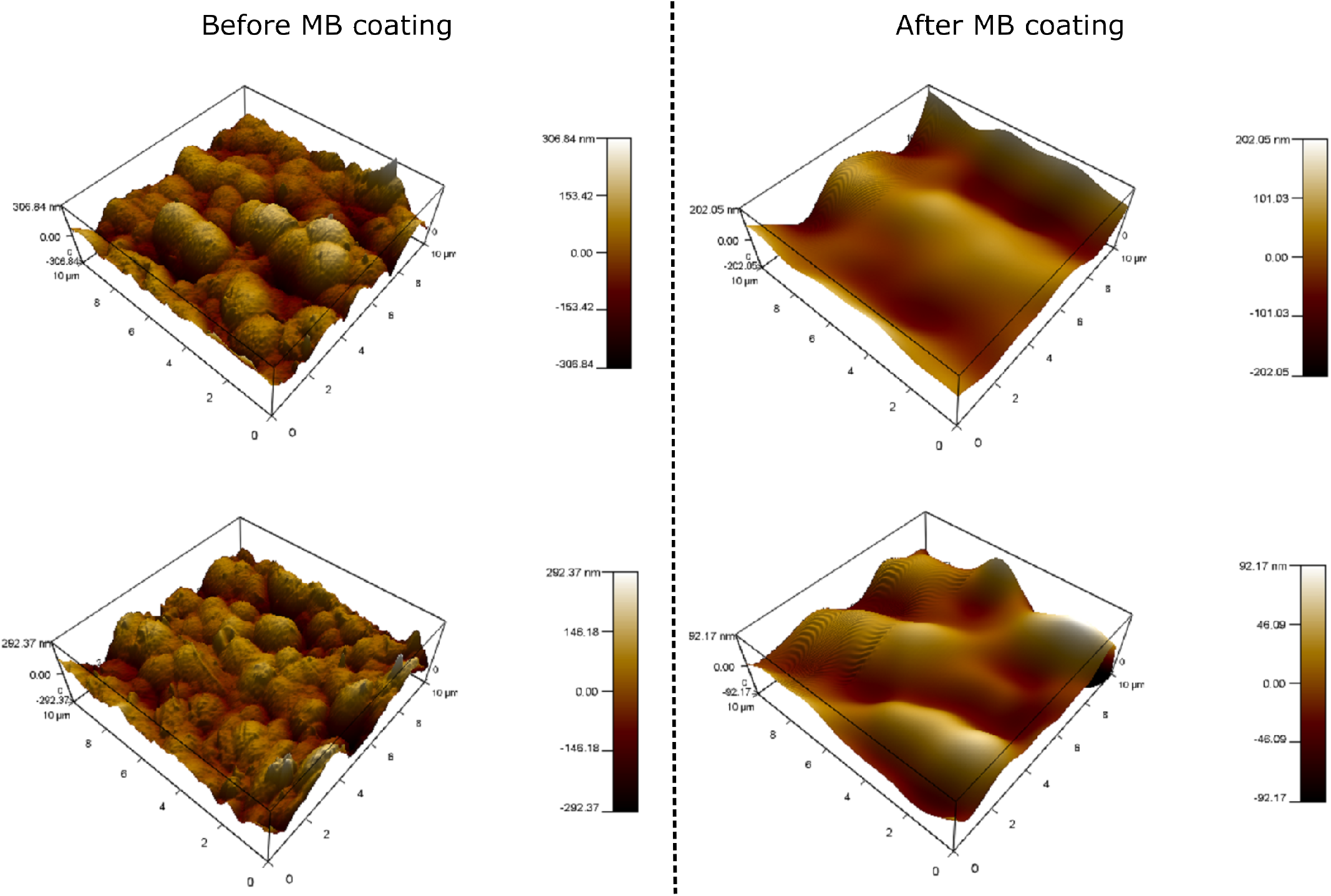
(a) AFM scanning results on PCB surface before after coating with methylene blue.

**FIG. S4.**
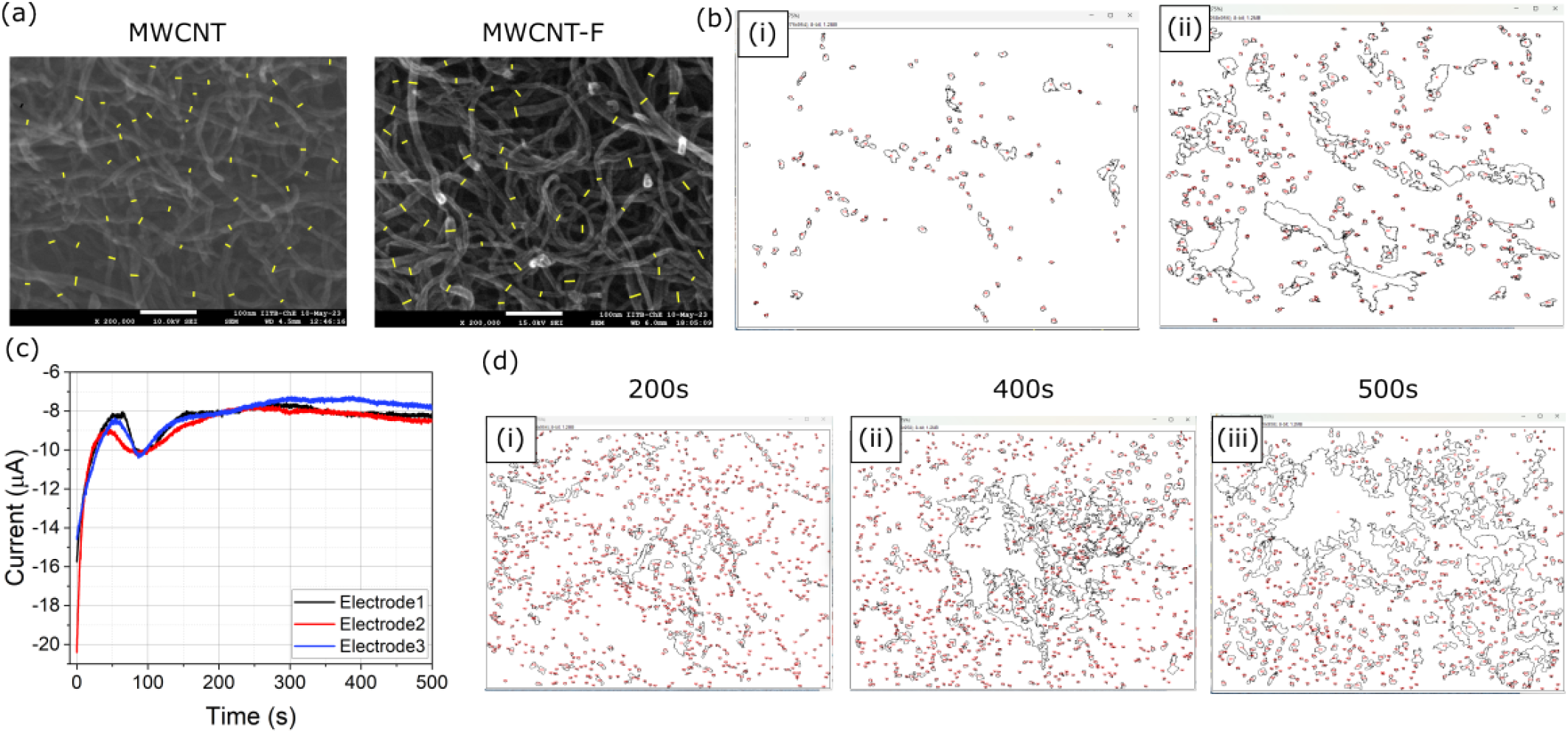
(a) SEM micrographs of MWCNT and MWCNT-F annotated in ImageJ software to analyze change in diameter of MWCNT due to acid functionalization. (b) ImageJ marked outline image of AuNPs grown on (i) MWCNT and (ii) MWCNT-F. MWCNT-F sample has more AuNPs than MWCNT. (c) Chronoamperometry profile for electrodeposition of AuNP on 3 PCB electrodes. (d) ImageJ marked outline image of AuNPs electrodeposited for varying chronoamperometry duration: (i) 200 s, (ii) 400 s, and (iii) 500 s. AuNPs deposited via chronoamperometry for 500 s are larger in size and form clusters.

**FIG. S5.**
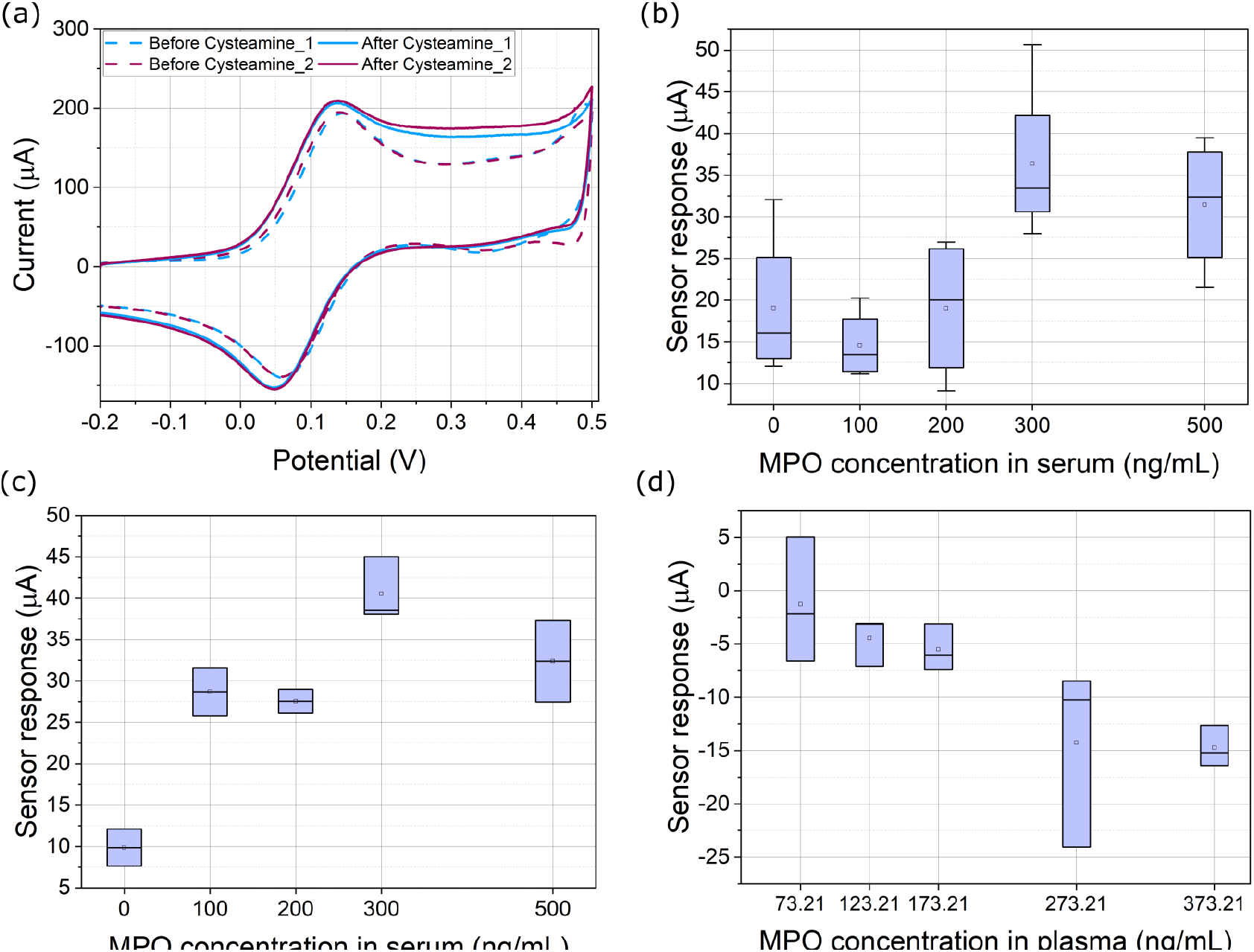
(a) CV voltammograms on 2 PCB electrode before and after coating cysteamine on AuNPs. After coating cysteamine, the CV peak current increases slightly, confirming coating of cysteamine. (b) Sensor response for various concentrations of MPO spiked in commercially procured human serum samples, on PCB electrodes without microfluidic packaging. (c) Sensor response for various concentrations of MPO spiked in commercially procured human serum, on PCB electrodes with microfluidic packaging. (d) Sensor response for various concentrations of MPO spiked in plasma extracted from human venous blood sample.

**FIG. S6.**
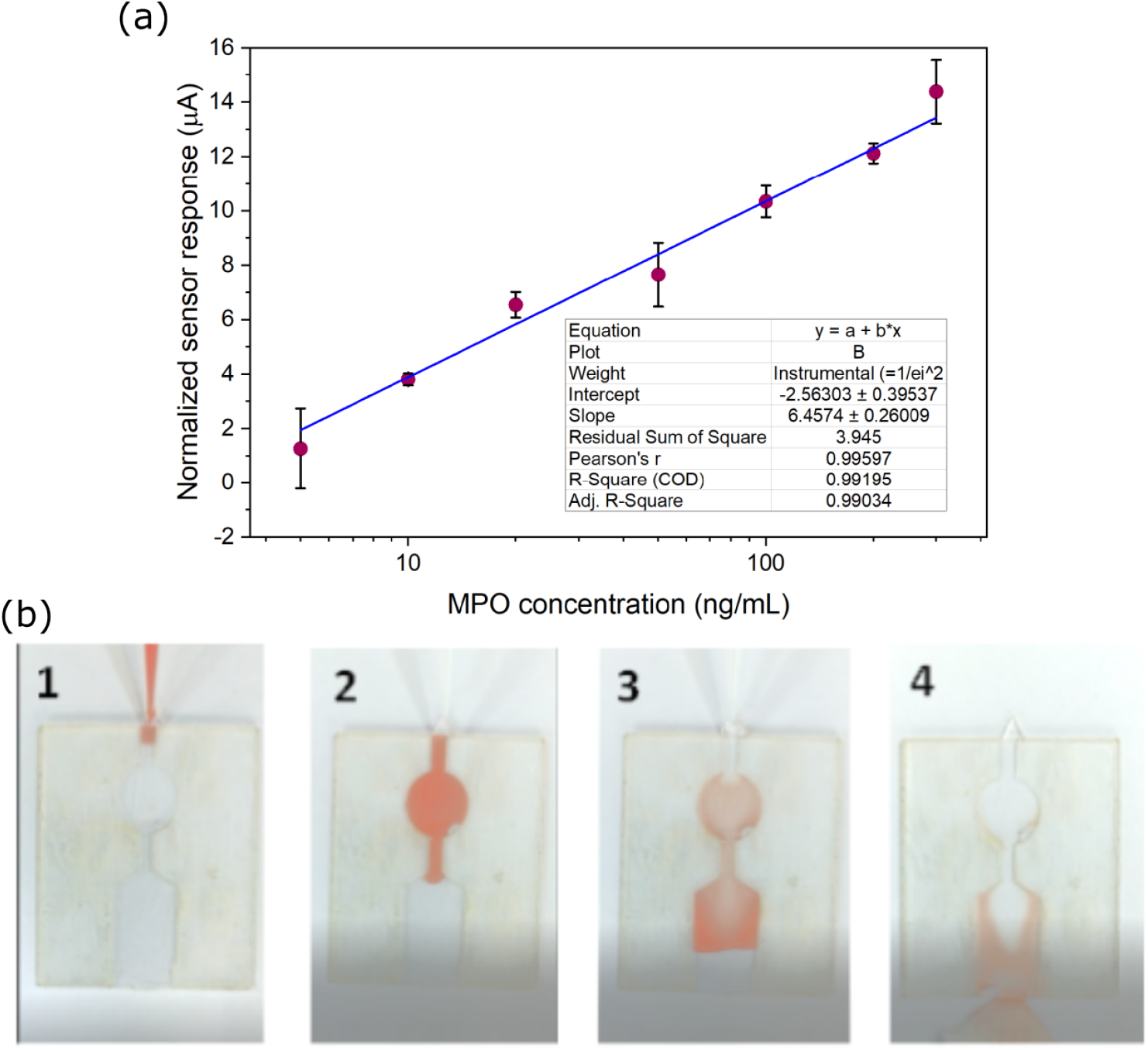
(a) Dose-response curve obtained with various MPO concentrations spiked in PBS. Relevant regression parameters are shown in the figure. (b) Microfluidic design prototype made with laser-cut double-sided tape encapsulated with acrylic sheets, tested using diluted red paint to show steps involved in sample testing: (1) dispensing 30 μL of sample, (2) sample covers the testing region, (3) Sample is pushed out of the testing region by introducing the next chemical (in this case, DI water), (4) sample is completely washed away from the testing region and absorbed using lint-free paper.

